# A novel, high throughput, and low-cost method for the detection of 40 amines relevant to inborn errors of metabolism, in under 60 minutes, using reverse phase high performance liquid chromatography

**DOI:** 10.1101/2024.05.09.24306940

**Authors:** Kirkland A Wilson, Yun Zhou, Gary Cunningham, Kimberly Chapman, Marshall Summar, Debra Regier

**Affiliations:** Children’s National Rare Disease Institute, Children’s National, Washington, DC 20012; Genetics and Molecular Biology Branch, NHGRI, NIH, Bethesda, MD 20892; Uncommon cures, LLC, Chevy Chase, MD 20815

**Keywords:** Amino Acid, Amine, RP-HPLC, Chromatography, Metabolomic, IEM

## Abstract

**Objectives:** An assessment of amino acid and amine concentrations is important for the diagnosis and management of inborn errors of metabolism (IEMs). Methods exist that measure these biologically important metabolites but are cost-prohibitive and/or time consuming. We therefore sought to develop a novel methodology, applicable to IEMs, that is both high-throughput and low cost.

**Methods:** Previously, we developed a methodology for rapid, repeatable, and cost-efficient separation of approximately 20 amines as a proof of concept and now expand it to amines relevant to IEMs. We describe our separation methodology using reverse phase high performance liquid chromatography with ultraviolet-visible spectrum absorbance paired with precolumn derivatization with *o*-phthaladehyde.

**Results:** We show reproducibility via concentration assessments, in triplicate, for each amine. We assess amines in prepared standard solutions and in biologic samples from patients with IEMs. We also detected and assessed the amino group containing compounds oxidized and reduced glutathione and ammonia. Validation was established using absolute area under the curve (AUC) and via comparison using a single internal standard.

**Conclusions:** We report good separation of 40 primary amino group containing metabolites, in a single, less than 60-minute run. This rapid, low cost, and accurate methodology only requires a small volume of sample and can greatly increase availability and access. Finally, the numerous disease associated amines (ie homocitrulline, trimethyllysine, alloisoleucine) and unique compounds detected in our single run has broad research and clinical utility and can increase efficiency, important as the need for analysis of amines grows globally.

## Introduction

Amino acids are vital metabolites that are used by the body to form proteins and neurotransmitters, they function in epigenetic regulation, act as an energy source, and have many other critical functions in health and disease. Given the many roles for amino acids, a large number of inherited and acquired disorders perturb amino acid concentrations including many inborn errors of metabolism (IEMs), insulin resistance, depression, organ dysfunction, and cancer.(1–4) Indeed, we have previously published on the critical role of amino acid homeostasis in sickle cell crisis.(5) Analysis of amino acids is critical in not only diagnosis, but also prognosis and clinical monitoring in many disorders.(6)

Ion exchange technologies with post-column derivatization using ninhydrin is the classic methodology for amino acid analysis and it is often used clinically for amino acid separation. These protocols, however, suffer from long separation times and require dedicated equipment.(7) Similarly, pairing high performance liquid chromatography (HPLC) with mass spectrometry has gained popularity but it is too expensive for widespread, routine use, especially in lower resource communities.(8) Thus the development of analytical techniques with lower cost technologies have recently gained popularity.

Many techniques have been developed for the detection of amino acids on HPLC systems, including some techniques to use *o-*pthalaldehyde (OPA).(4,9–12) We have also published previously on the use of OPA for limited, 20 amino acid analyses and showed 99% concordance with the ninhydrin assay.(13) However, to our knowledge no methods exist to date that allow for rapid separation of greater than 20 amino group containing metabolites, known as amines, nor for those amines specifically relevant to IEMs, in human biologic samples using a single injection.

It is becoming increasing obvious that amine homeostasis plays a role in many disorders, both common and rare.(1–4) A barrier to the application of amine analysis to both research and clinical care, however, has been the high cost, time, and difficulty of current technologies. We therefore describe here a rapid, low-cost, and high throughput, less than 60-minute, protocol for the detection of 40 primary amino group containing metabolites. We furthermore tested our methodology in biologic samples and specifically in samples from patients with IEMs. We suggest that this protocol could allow for immediate application and the widespread assessment of clinically relevant compounds, especially in lower resource environments. This will allow for the widespread application of amine analysis and the ease of the methodology will allow for its immediate use. This work expands on our previous peer reviewed methodology in an attempt to increase access to an amine analysis.(13)

## Materials and Methods

### Chemicals and columns

The Infinity Lab Poroshell 120, 2.7 μm C18 analytical column and the corresponding guard column, borate buffer and OPA reagent were purchased from Agilent Technologies (Santa Clara, CA). HPLC grade acetonitrile, methanol, and water were purchased from VWR International (Radnor, PA). Individual amino acids and D-α-aminobutyric acid (AABA), the internal standard, and all other chemicals were purchased from Sigma Aldrich (St. Louis, MO). Argon gas was purchased from Roberts Oxygen Co (Rockville, MD).

10 mM stocks of the individual amino acids were prepared separately and stored at -80 °C until use. To check linearity and reproducibility, concentrations from 0 to 2500 μM were made by serial dilutions. Preparations were thawed or prepared fresh on the day of analysis, except for the 4 °C storage variability test, termed the delayed sample analysis variability test, which was conducted with the same sample over three days. Each amino group containing metabolite was run independently and in aggregate to both resolve co-migration and to verify retention time.

### Equipment

The 1290 Infinity II LC System was purchased from Agilent Technologies (Santa Clara, CA), with the addition of a 40 μL syringe and sample loop for the HPLC system. The Centrifuge 5417c was purchased from Eppendorf (Hamburg, Germany). The vortex mixer was purchased from BioExpress (Kaysville, UT). The 3k centrifuge filters were purchased from VWR international (Radnor, PA).

### Chromatographic conditions

Derivatization and injection are as described in our prior paper.(13) A binary mobile phase consisting of solution A, 20 mM sodium phosphate (dibasic), 20 mM sodium borate, and 5 mM sodium azide, pH adjusted to 7.2, and solution B, a mixture of 45% acetonitrile, 45% methanol, and 10% water were used. Programming for the chromatographic run starts with 98% of solvent A, 2% solvent B and using multiple isocratic stepwise increases (Table 1) over a 41 minute time course reduces A to 40% and B to 60%. This is termed the Isocratic Elution Phase. A final column elution step using 80% solution B and 20% water, solution C is then run over 4 minutes. This is termed the Carryover Reduction Phase. Finally, the column was set back to 98% solvent A and 2% solvent B to re-equilibrate over 4 min. This is termed the Re-equilibration phase. The total run time is 53 minutes.

**Table 1.** Concentration of Mobile Phase. Analyte elution occurs through a series of isocratic steps with increases of the organic mobile phase buffer. Solution A, the non-organic buffer, is 20 mM sodium phosphate (dibasic), 20 mM sodium borate, and 5 mM sodium azide, adjusted to pH 7.2. Solution B, the organic buffer, is a mixture of 45% acetonitrile, 45% methanol, and 10% water. Solution C is water.

| Time (minutes) | Solution A (%) | Solution B (%) | Solution C (%) |
| --- | --- | --- | --- |
| 0 | 98 | 2 | 0 |
| 4 | 87.7 | 12.3 | 0 |
| 15 | 87.7 | 12.3 | 0 |
| 16 | 79 | 21 | 0 |
| 22 | 79 | 21 | 0 |
| 23 | 75 | 25 | 0 |
| 27 | 75 | 25 | 0 |
| 27.5 | 72 | 28 | 0 |
| 31 | 72 | 28 | 0 |
| 32 | 64 | 36 | 0 |
| 38 | 64 | 36 | 0 |
| 41 | 40 | 60 | 0 |
| 43 | 0 | 80 | 20 |
| 47 | 0 | 80 | 20 |
| 49 | 98 | 2 | 0 |
| 53 | 98 | 2 | 0 |

The column temperature was held constant at 34 °C while the sample tray was maintained at 4 °C. UV detection was performed at 338 nm.

### Concentration Curve

A low concentration curve (0, 1, 2.5, 5, 10, 20, 30, 40, 100, 500, 1000, 1500, and 2500 μM) and high concentration curve (0, 5, 15, 20, 30, 40, 100, 150, 200, 500, 1000, 1500, and 2500 μM) were used to assess coverage across both normal and abnormal concentrations. These standard curves, conducted in triplicate, were based on published reference ranges and were used to evaluate the analytical measurement range.(14)

### Method Validation

Table 2 reports the methodology validation characteristics for each compound. All metrics were conducted in triplicate. Baseline noise was calculated on a per compound basis by assessing the baseline signal intensities at the expected retention time for the 0 μM time point, averaged, and calculated based on the representative concentration for the analyte. The limit of detection (LoD) for each compound was calculated based on the agreed upon International Committee on Harmonization and the Clinical and Laboratory Standards Institute formula, LoD = 3.3*S/s.(15,16) Where S is the variation of the background noise and s is the slope of the concentration curve for that compound. Limit of quantitation (LoQ) was calculated based on a 10:1 ratio as compared to the LoD. The carryover was determined by running a blank sample, after each 2500 μM sample in the concentration curves and assessing for signal intensity at the relevant retention time. Intra-assay variability was assessed via assay of the same sample during the same sequence as subsequent runs. Inter-assay variability was assessed via separate assays as different run sequences. To additionally interrogate sample stability during a large sample run, a delayed sample analysis variability test, where samples are left in the HPLC sample tray at 4 °C for an extended period of time, a single sample was left in the HPLC sample tray at 4 °C for a full 48 hours and assessed initially and then at 24 hour intervals, the “4 °C Storage Variability” in Table 2.

**Table 2.** Method validation parameters per analyte. All measurements are reported in μM. Metrics are reported based on calculations at the expected retention time per analyte. Variability assessments are based on 100 μM analyte concentrations.

|  | Compound | Baseline Noise | Limit of Detection | Limit of Quantitation | Carryover | Intra-assay Variability | Inter-assay Variability | 4 °C Storage Variability |
| --- | --- | --- | --- | --- | --- | --- | --- | --- |
| 1 | 1-Methylhistidine | 0.85 | 1.30 | 3.94 | $2.80 \pm 0.30$ | 0.12 | 0.51 | 1.62 |
| 2 | 3-Methylhistidine | 0.44 | 0.62 | 1.89 | $3.35 \pm 0.49$ | 0.11 | 0.51 | 1.65 |
| 3 | Alanine | 1.82 | 1.92 | 5.82 | $7.78 \pm 5.38$ | 0.20 | 0.83 | 0.10 |
| 4 | Alloisoleucine | 0.95 | 1.98 | 5.99 | $1.97 \pm 0.37$ | 0.17 | 0.42 | 3.43 |
| 5 | Ammonia | 0.91 | 9.69 | 29.35 | $0.18 \pm 0.06$ | 0.30 | 0.55 | 12.07 |
| 6 | Anserine | 2.94 | 4.16 | 12.62 | $8.04 \pm 4.45$ | 0.13 | 1.05 | 2.52 |
| 7 | Arginine | 0.73 | 0.97 | 2.95 | $3.70 \pm 0.19$ | 0.10 | 0.95 | 2.53 |
| 8 | Argininosuccinate | 0.11 | 0.58 | 1.75 | $0.13 \pm 0.001$ | 0.18 | 0.55 | 8.32 |
| 9 | Asparagine | 0.39 | 0.55 | 1.68 | $2.65 \pm 1.24$ | 0.42 | 1.11 | 2.86 |
| 10 | Aspartate | 0.62 | 0.85 | 2.58 | $2.72 \pm 0.43$ | 1.05 | 0.53 | 0.12 |
| 11 | Beta-alanine | 0.24 | 0.28 | 0.83 | $0.69 \pm 0.2$ | 2.22 | 0.97 | 1.47 |
| 12 | Carnosine | 1.12 | 1.58 | 4.78 | $2.09 \pm 1.08$ | 0.28 | 1.37 | 4.96 |
| 13 | Citrulline | 0.59 | 0.73 | 2.22 | $1.43 \pm 0.57$ | 0.12 | 1.18 | 2.28 |
| 14 | Cystathionine | 1.63 | 1.33 | 4.02 | $7.05 \pm 1.83$ | 0.09 | 1.26 | 1.54 |
| 15 | Cystine | 0.60 | 0.65 | 1.96 | $2.46 \pm 0.61$ | 0.23 | 0.46 | 2.90 |
| 16 | Ethanolamine | 2.45 | 2.10 | 6.36 | $29.36 \pm 19.65$ | 0.42 | 0.44 | 1.45 |
| 17 | γ-Aminobutyric Acid (GABA) | 0.90 | 1.68 | 5.09 | $0.99 \pm 0.73$ | 0.27 | 1.27 | 3.47 |
| 18 | Glutamate | 0.88 | 1.09 | 3.31 | $3.78 \pm 1.43$ | 0.13 | 1.57 | 2.04 |
| 19 | Glutamine | 0.99 | 1.21 | 3.67 | $6.45 \pm 3.63$ | 0.16 | 2.28 | 1.89 |
| 20 | Glutathione (GSH) | 0.26 | 4.24 | 12.83 | $0.03 \pm 0.01$ | 3.92 | 2.81 | 14.96 |
| 21 | Glutathione Disulfide (GSSG) | 1.08 | 1.06 | 3.22 | $3.61 \pm 1.10$ | 1.12 | 1.18 | 1.62 |
| 22 | Glycine | 1.46 | 2.96 | 8.96 | $4.06 \pm 0.60$ | 1.94 | 0.66 | 7.81 |
| 23 | Histidine | 0.47 | 1.35 | 4.09 | $1.12 \pm 0.65$ | 0.63 | 0.25 | 7.60 |
| 24 | Homocitrulline | 0.42 | 0.54 | 1.62 | $3.48 \pm 3.44$ | 0.35 | 1.87 | 2.47 |
| 25 | Homocysteine | NOT QUANTIFIABLE |  |  |  |  |  |  |
| 26 | Homocystine | 0.63 | 0.40 | 1.21 | $8.13 \pm 0.44$ | 0.18 | 1.39 | 2.17 |
| 27 | Isoleucine | 2.28 | 2.87 | 8.68 | 6.52 ± 0.15 | 0.48 | 1.08 | 2.12 |
| 28 | Leucine | 1.60 | 1.99 | 6.02 | 4.54 ± 0.51 | 0.71 | 0.90 | 8.31 |
| 29 | Lysine | 0.47 | 0.45 | 1.35 | 11.35 ± 0.42 | 0.43 | 0.92 | 7.54 |
| 30 | Methionine | 0.78 | 1.01 | 3.06 | 1.84 ± 0.32 | 0.17 | 0.65 | 1.58 |
| 31 | Ornithine | 0.87 | 0.80 | 2.42 | 7.15 ± 1.02 | 0.16 | 1.08 | 0.97 |
| 32 | Phenylalanine | 0.56 | 0.70 | 2.12 | 4.44 ± 4.20 | 0.13 | 0.62 | 2.23 |
| 33 | Serine | 1.26 | 1.75 | 5.31 | 3.96 ± 0.45 | 0.21 | 1.41 | 4.43 |
| 34 | Sulfocysteine | 0.06 | 0.07 | 0.20 | 1.00 ± 0.10 | 0.60 | 0.58 | 2.15 |
| 35 | Taurine | 0.30 | 0.40 | 1.23 | 1.45 ± 0.65 | 1.80 | 0.20 | 6.92 |
| 36 | Threonine | 1.16 | 1.51 | 4.58 | 3.99 ± 1.08 | 0.36 | 0.57 | 1.49 |
| 37 | Trimethyllysine | 0.69 | 1.02 | 3.09 | 2.85 ± 0.60 | 0.33 | 0.29 | 10.26 |
| 38 | Tryptophan | 0.26 | 0.35 | 1.06 | 0.92 ± 0.71 | 1.69 | 0.59 | 18.36 |
| 39 | Tyrosine | 0.25 | 0.33 | 1.01 | 1.22 ± 0.41 | 0.38 | 0.89 | 7.79 |
| 40 | Valine | 1.38 | 1.81 | 5.48 | 2.47 ± 2.57 | 0.43 | 0.46 | 2.57 |

### Biologic Samples

Biologic samples as plasma or whole blood were obtained from healthy patients and those with different IEMs and either processed immediately or stored at -80°C until use. Samples were processed as previously described.(13) Patient and healthy participants provided informed consent under the study protocol (CN PRO0004911). Samples were processed and analyzed in a blinded fashion, without knowing the underlying disease status prior to analysis.

## Results

### Ideal Chromatographic Conditions (Table 1, Figure 1)

Chromatographic conditions for optimal separation with minimal tailing were determined by preparing and injecting amino acids independently and in aggregate to resolve co-migration. Adjusting either the concentration of the mobile phase and/or the column temperature were the conditions found to have greatest effect on separation. Column temperature affected the entire chromatography spectra and was less sensitive for specific amino acid resolution than variation in mobile phase solutions but the column temperature for optimal baseline separation was determined to be 34°C. Optimal chromatographic mobile phase adjustments were found to display better resolution using isocratic stepwise changes for resolution rather than using a constant or variable gradient elution chromatographic condition. The vast majority of compounds elute during these isocratic holds rather than during the gradient increases between the isocratic holds (Figure 1).

**Figure 1.**
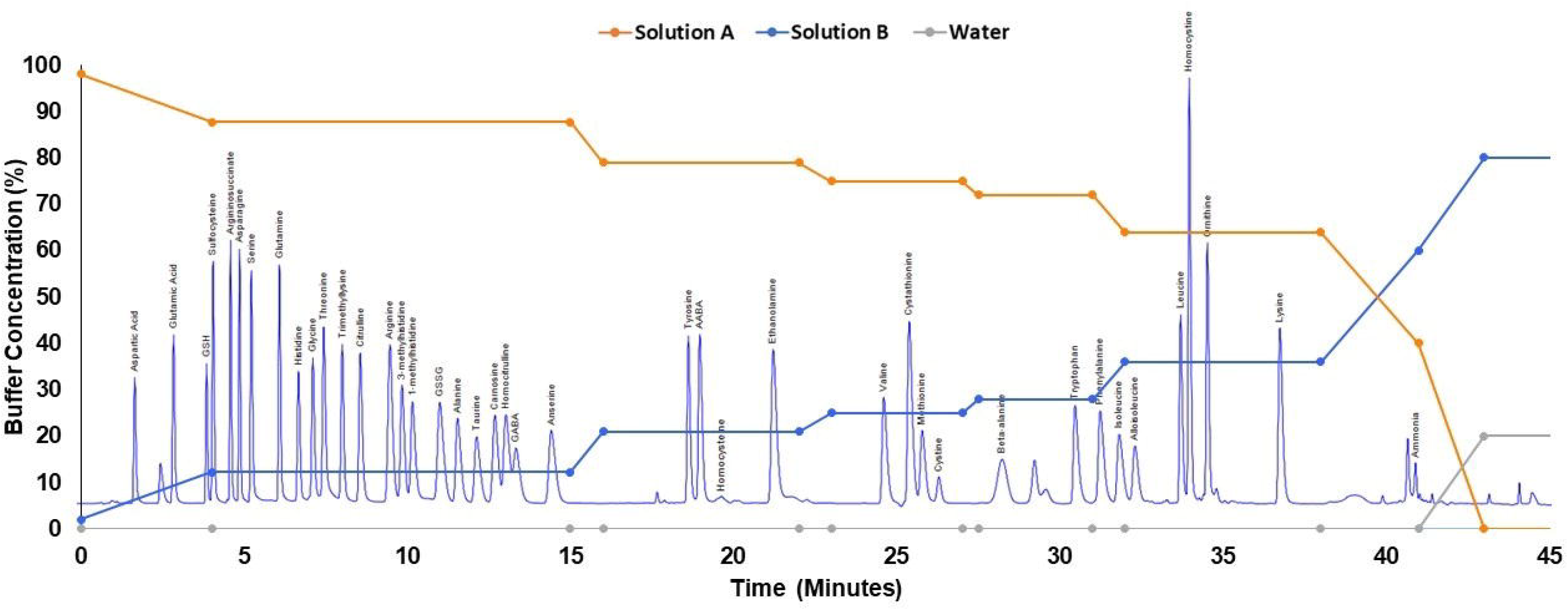
Chromatograph Showing Separation of Amine Standards. All metabolites were run at 100μM. Unlabeled peaks are not unique to any particular metabolite when metabolites are run separately. Overlaid on the chromatograph is the buffer gradient changes.

Unlabeled peaks are non-specific peaks that are not unique to any particular amine and instead are likely due to the derivative agent, OPA, when seen in the standards.

We found that carryover of samples was minimized by the addition of a Carryover Reduction Phase to the chromatographic conditions. Specifically, carryover was maximally minimized with a mixture of 80% of solution B with 20% water. In order to reduce variation in analyte retention time, a Re-Equilibration Phase was added to the end of the run, prior to injection of the next sample. Four minutes was found to be the minimal amount of time necessary to allow for reequilibration.

Concentration curves using both absolute metabolite area under the curve (AUC) and a ratio of the metabolite AUC to internal standard AUC show good correlation for all analytes. Linear fit of all amines was assessed using linear regression and was R^2^ ≥ 0.99 for all compounds (Data not shown).

### Method Validation Parameters (Table 2)

We assessed the method validation parameters for each of the 40 analytes. Baseline noise, which was directly measured as signal intensity and converted to assumed concentrations of interference per amine, was less than 3 μM for all analytes. The chromatographic conditions gave a LoD less than 5 μM for all compounds except for ammonia. Similarly, the LoQ, was the worst for ammonia. For carryover, only ethanolamine, after a 2.5 mM concentration sample, had sample carryover greater than 12.5 μM or 0.5% of the prior sample concentration. Specifically, ethanolamine showed an average carryover of 1.17% of the maximal concentration of 2.5 mM. Measures of precision were conducted for the method as well and intra-assay and inter-assay variability was less than 4 μM, or 4% of the known concentration, for all compounds including the labile compound, glutathione.

As we purport that this method can increase the access to an amino acid analysis and therefore increase the number of analyses run sequentially, we conducted an additional measure of variation to assess the amount of variance that occurs between samples when stored in the sample tray, which is maintained at 4 °C in our instrument. This would mimic a long sample run in which they are left for 24 hours or more after preparation, such as to run overnight. As expected both ammonia and glutathione showed greater than 10% variation from the known concentration in this delayed sample anlaysis variability test, as did trimethyllysine and tryptophan.

Cysteine was not well detected via this methodology. A concentration curve was run for this compound and poor resolution from the baseline was noted for concentrations under 500 μM. Additionally, a second peak in the concentration curve generated for cysteine was noted to have a linear increase in concentration with cysteine concentration. Based on retention time this was determined to be cystine that resulted from the well-known spontaneous oxidation reaction of cysteine to the disulfide, cystine, at neutral pH.(17,18) Thus, in our method, the peak representing cystine was deemed to be a combined cysteine + cystine peak. Homocysteine was also unstable via this method, and spontaneously formed its disulfide homocystine. While homocysteine could be qualitatively observed and resolved from other peaks, unlike cysteine, but only on samples run within 24 hours of preparation, it could not be reliably quantitated (Figure 1). Also, there was no significant effect on inter-assay variability for the disulfide compounds suggesting that the vast majority of cysteine and homocysteine is very rapidly, before detection, oxidized to the disulfide form.

The OPA derivatization reaction was conducted in the presence of a thiol reducing reagent, specifically 2-mercaptoethanol. There was concern for multiple peaks from replacement of 2-mercaptoethanol by the thiol group in thiol containing amines, namely glutathione, cysteine, and homocysteine. Specifically, whether there would be a peak with OPA, the 2-mercaptoethanol thiol group, and the relevant amine, and a second peak without the 2-mercaptoethanol thiol group but instead with the sulfhydryl group internal to the amino acid. Notably, none of the thiol group containing amino acids gave this unique secondary peak.

### Application to Biologic Samples

Biologic samples from a patient without a documented IEM (Figure 2) as well as multiple patients with documented IEMs, specifically maple syrup urine disease (MSUD), ornithine transcarbamylase deficiency, and carbamoyl phosphate synthetase 1 deficiency, were assessed for practical application of the method. We furthermore wanted to determine the ability to resolve amines in both red blood cells and plasma. Biologic samples showed good separation without need for further adjustment for separation of unidentified peaks in both sample types. Using this methodology we were able to identify and quantitate all clinically relevant peaks, in a blinded fashion, that led to the correct clinical diagnosis. Notably, on prior analysis of the same sample from a patient with MSUD conducted by our clinical laboratory using a ninhydrin-based assay the pathognomonic compound, alloisoleucine, was only qualitatively reported via that technology.

**Figure 2.**
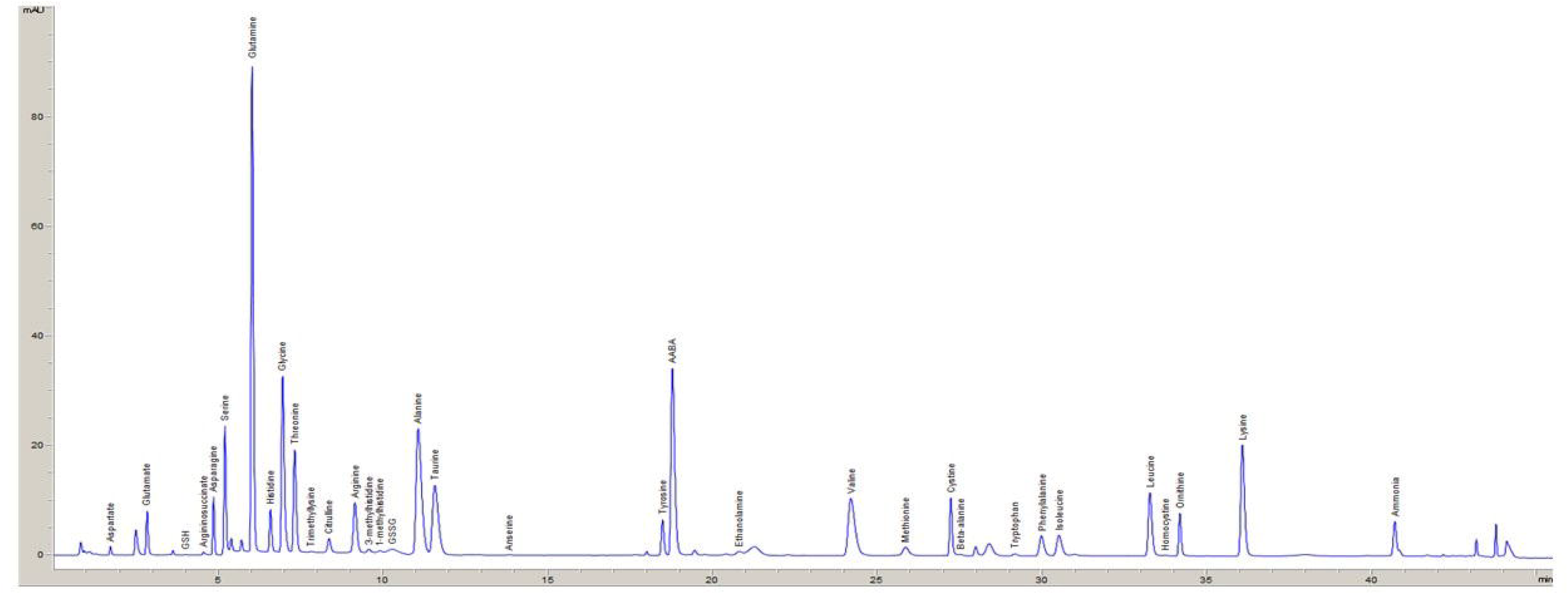
Plasma from a Biologic Sample. Shown is a sample from a healthy volunteer to represent how the methodology functions in a biologic matrix. Unlabeled peaks were unidentified and distinct, based on retention time, from known analytes based on metabolite standards run during the same chromatographic program.

## Discussion

This method represents a novel and practical approach to amine analysis for the largest number of amino group-containing compounds to date. It is not limited solely to amino acids, but can be applied to any primary amino group containing compounds, making it a very expansive technique. Additionally, we only require a single injection for resolution of 40 analytes and we are able to separate and quantitate a number of compounds that classically have poor or no resolution on other technologies such as alloisoleucine and cystathionine, and argininosuccinate and leucine.(12,19)

Baseline resolution is achieved for the vast majority of compounds while still allowing for a less than 1 hour total runtime. While ethanolamine had the largest carryover of all the analytes, it should be noted that it was less than 2% of the massively supraphysiologic concentration used, 2.5 mM. For reference, the normal concentration of ethanolamine in blood is 2-12 μM.(20) Given the large concentration required to reach this small amount of carryover, it is expected to have minimal effect in nearly all cases.

We noted that the LoD and LoQ was highest for ammonia and while the intra-assay and interassay variability were both excellent the “real-world” 4 °C storage variability, which we used to simulation a long multi-sample run, showed the expected elevation in variability in ammonia concentration. We suspect that this increased variability is due to the known spontaneous deamination that can occur in samples and this was an expected limitation of the methodology.(21) We do not suggest that this method should replace standard ammonia assessment techniques but instead only note that we can also assess this analyte via our methodology. Separately, we note that for glutathione, tryptophan, and trimethyllysine, the delayed sample analysis variability was also greater than 10μM, or 10% of the analyte concentration, suggesting that per our method these compounds are not stable for an extended time after preparation, when left in the HPLC system at 4 °C. Thus, samples in which any or all of these analytes are of concern should be analyzed within 24 hours of sample preparation, or frozen until ready for analysis.

We were not able to accurately quantify cysteine nor homocysteine in our methodology. Given that our method is run at neutral pH, which is known to cause oxidation of these compounds to their disulfide forms, we suspect that an adjustment to an acidic pH would improve detection of these compounds.(17,18) Further study is needed to assess for ideal conditions for these analytes and the effects of a change in pH on the remaining analytes.

Our improved method allows for the separation of 40 primary amino group containing metabolites and was applied to biologic samples in patients with disorders that result in disruptions in amine concentrations. We show minimal carryover at high concentrations and good reproducibility as inter-run and inter-day assessments. The linear regressions of our analytes show excellent precision. We assessed this methodology in a blinding fashion and accurately identified and quantitated the relevant compounds and disease states without prior knowledge of the diagnosis. Given the diagnostic and prognostic significance of amines, our methodology, which shows utility in biologic fluids, provides an immediate solution to expanding access to those communities that cannot utilize the other methodologies for an amine analysis. This fulfills a large unmet clinical need as the Genetic Metabolic Dieticians International organization cites the limitations of amino acid technologies and access as barriers to implementation of their guidelines for monitoring in IEMs.(22–24) This will also allow for further investigations, including use in a greater number of patients to assess the full range of clinical applicability of our methodology. Notably, as with our original methodology, we preserve the small volumes of sample needed, which can be critical in the newborn period and/or with patients who would require large volume blood draws for other analysis.

## Data Availability

All data produced in the present study are available upon reasonable request to the authors

## Abbreviations

Amino acid: (AA)
Area under the curve: (AUC)
High performance liquid chromatography: (HPLC)
Inborn error of metabolism: (IEM)
Maple syrup urine disease: (MSUD)
*o*-pthaldehyde: (OPA)

